# Adolescence is characterized by more sedentary behavior and less physical activity even among highly active forager-farmers

**DOI:** 10.1101/2023.03.15.23287308

**Authors:** Ann E. Caldwell, Daniel K. Cummings, Paul L. Hooper, Benjamin C. Trumble, Michael Gurven, Jonathan Stiegltz, Helen E. Davis, Hillard Kaplan

## Abstract

Over 80% of adolescents worldwide are insufficiently active, posing massive public health and economic challenges. Declining physical activity (PA) and sex differences in PA consistently accompany transitions from childhood to adulthood in post-industrialized populations and are attributed to psychosocial and environmental factors. An overarching evolutionary theoretical framework and data from pre-industrialized populations are lacking. In this cross-sectional study we test a hypothesis from life history theory, that adolescent PA reductions reflect an evolved strategy to conserve energy, given the increasing sex-specific energetic demands for growth and reproductive maturation. Detailed measures of PA and pubertal maturation are assessed among Tsimane forager-farmers (age: 7-22 yrs.; 50% female, n=110). We find that 71% of Tsimane sampled meet World Health Organization PA guidelines (≥60 minutes/day of moderate-to-vigorous PA). Consistent with post-industrialized populations, we observe sex differences and inverse age-activity associations mediated by Tanner stage. Physical inactivity in adolescence is distinct from other health risk behaviors and also not merely resulting from obesogenic environments.

## Introduction

Inadequate physical activity (PA) is one of the leading causes of global chronic disease burden, responsible for billions of dollars in healthcare costs and lost productivity each year [1]. Recent population-based surveys of self-reported PA found 28% of adults worldwide are insufficiently active, defined as less than150 minutes/week of moderate intensity PA or 75 minutes/week of vigorous PA [2]. Additional concern stems from the recognition that a staggering 80% of adolescents (11-17yo) worldwide are insufficiently active, defined as less than 60 minutes/day of moderate-to-vigorous PA [3]. True prevalence of inadequate PA is likely even higher because self-reports typically overestimate PA relative to wearable motion devices [4–6]. In the US, just 5% of the adults and 8% of adolescents meet PA guidelines when measured via actigraphy.

Two of the most consistent findings in epidemiological studies are that PA declines with age and that males are more active than females [7,8]. The steepest age-related declines in PA are observed during adolescence and occur at earlier ages for girls than boys in several post-industrialized countries [5,7,9–14]. A pooled analysis of 26 longitudinal studies of primarily self-reported PA in adolescents aged 10-19 y estimates a 7% decline in PA per year [9]. Long-term longitudinal studies of adolescents in the UK and Canada found that low PA levels following these precipitous declines largely persist into adulthood [13,14], a pattern also supported by cross-sectional epidemiological research in the US [5]. It is critical to understand factors underlying these consistently observed sex differences and decreases in PA in adolescence to inform the development of more targeted and effective PA interventions.

Although epidemiological patterns of PA in adolescence are consistent across most industrialized populations studied to date, we have a limited understanding of factors influencing PA during adolescence in settings more reminiscent of our pre-industrial past. While no single population represents the mosaic of environments in which humans evolved, populations living a subsistence-based lifestyle that necessitates high levels of obligatory PA, with little to no access to social media, television, or video games offer interesting contrasts to industrialized populations characterized heavily by more sedentary lifestyles in urbanized settings. Research on PA in adolescents in these populations is needed to help disentangle the influence of post-industrialized environments and lifestyle versus developmental constraints on PA during the transition from childhood to adulthood.

A growing body of epidemiological research shows that PA during adolescence is not only associated with age, but also with timing of biological maturation, and sex differences in the pace of PA decline are attenuated when using an estimate of maturation-relative age (i.e., biological age), rather than chronological age [7,14–21]. When in the same statistical model, maturation-relative age more strongly predicts PA than chronological age [7]. A range of measures have been used to estimate biological age in these studies including predictive equations to estimate proportion of adult height reached [17] or age from peak height velocity based on population-based averages [7,19], while a few studies have measured peak height velocity [14,22] and/or used self-reported measures of Tanner stage of reproductive maturation [14,21]. Notably, biological age is often considered a factor that “confounds” the understanding of sex differences and age-related declines in PA [e.g., 15,17,19].

One hypothesis to explain the association between biological maturation and PA is that the psychosocial stress of puberty leads adolescents to adopt ‘health risk’ behaviors (e.g., tobacco, alcohol, and drug use, and unprotected sex) [13,15]. Physical inactivity is considered a health risk behavior in post-industrialized settings, where the leading causes of death are non-communicable diseases (e.g., heart disease, stroke, diabetes), inactivity is pervasive, and PA occurs primarily through leisure time exercise rather than through obligatory subsistence labor or transportation. To further test predictions based on this and other hypotheses, studies are needed that include objective measures of PA, more accurate measures of biological maturation, and are conducted in populations where physical inactivity is not necessarily a health risk behavior.

Life history theory provides an alternative framework to derive hypotheses for examining the relationships between age, biological maturation, and PA/inactivity, positing that because energy and time are finite, organisms face trade-offs in how to allocate time and energy during their lifespans between competing demands of growth, reproduction, and maintenance (i.e., immune system and somatic repair) [23,24]. The optimal way to allocate limited time and energy between these competing demands has been shaped by natural selection in ways that reliably promoted survival and reproductive success over large evolutionary time scales. Peter Ellison recently extended traditional life history models to integrate how energy intake and expenditure, including PA, also influence and are influenced by the primary life history trade-offs [25].

Hormones are key regulators that coordinate the strategic allocation of energy between somatic functions in ways that are age-, sex-, and environment-specific. Adolescence is a life stage characterized by distinct endocrinological, anatomical, and cognitive changes that likely require substantial energetic resources, though the energetic costs of these hormones and related physiological sequelae have not been fully quantified [26]. Puberty is associated with higher resting and total energy expenditure, partly due to increases in body size and fat-free mass; but skeletal growth, pubertal hormones, and neurocognitive changes are also assumed to play a role [26]. During periods of greater energetic demands for growth and development (i.e., adolescence, pregnancy, lactation), PA may be reduced, and sedentary time increased to help conserve energy for meeting these demands. Among Hiwi and Ache hunter-gatherers, for example, nursing mothers spend less time foraging and acquire less food than do non-nursing women [27].

A large body of research has demonstrated that high levels of PA, or low food intake, can also delay puberty, or suppress the female reproductive axis - adaptively shifting energy toward survival and away from reproduction when food availability is low or physical demands are high [28–36]. Sex differences in PA and sedentary time may therefore emerge regardless of environmental context during the transition from childhood to adulthood because sexually dimorphic changes prior to and during pubertal maturation increase the relative energetic costs of performing PA for females (e.g., smaller lung volume and capacity, lower cardiorespiratory fitness, lower anaerobic energy production and power, more body fat relative to fat-free mass) [37], while female reproductive maturation and fecundity can benefit from conserving energy and accumulating adipose tissue needed for pregnancy and lactation. In contrast, PA facilitates accumulation of muscle mass and strength, which is particularly beneficial for males in preparation for future adult reproductive and economic roles in environments with high levels of obligatory PA to subsist and survive, as was the case for the vast majority of human evolutionary history.

Examining PA in children and adolescents in non-industrialized environments more reminiscent of our evolutionary past can help clarify the roles of biology and environment for influencing sex differences in PA, and the relationship between biological maturation and PA. Toward that end, we conducted a cross-sectional study among Tsimane children and adolescents that included measures of PA time and intensity using Actigraph wGT3X+ accelerometers, contextualized with concurrent 24-hour PA recall interviews. We also assessed Tanner stage validated with hormonal and somatic measures of growth and maturation: dehydroepiandrosterone (DHEA), testosterone, height velocity, body fat, and grip strength. The Tsimane are an indigenous population of forager-horticulturalists residing in the Amazon basin in lowland Bolivia. They have little to no access to electricity or running water, show high levels of habitual physical activity [38], minimal hypertension (3.9%) [39], and the lowest recorded prevalence of coronary artery disease [40]. The vast majority of calories in the diet are produced through slash-and-burn horticulture, hunting, and fishing; <10% of calories are derived from store-bought goods (e.g. cooking oil, refined sugar, and salt) although market items are becoming increasingly prevalent [41].

Our overarching hypothesis is that the energetic needs for growth, reproductive maturation, and reproduction are heightened during adolescence and pose fundamental constraints on the energy available for PA across all environments. A negative association between age and PA is consistently observed during the transition from childhood to adulthood because these energetic demands trade-off against energy available for PA. Thus, we predict that age-activity associations would be observed among Tsimane adolescents, even though high levels of PA are necessary for daily life, and physical inactivity is not a health risk behavior in this population (P1). We also predict that Tsimane males would have higher levels of PA, similar to the sex differences observed in post-industrialized countries, because changes prior to and during reproductive maturation also change the costs and benefits of PA, favoring higher levels of PA in males (P2). Finally, we predicted that the association between age and PA is mediated by Tanner stage of reproductive maturation, supporting the hypothesis that the wide range of anatomical and neuroendocrine changes encompassed within Tanner stages explain the associations between age and PA, rather than confound them (P3).

## Results

### Sedentary Time and Physical Activity

Tsimane children and adolescents spent 356±80 minutes of daytime hours in sedentary time, 327±76 minutes/day in light intensity PA, 92±49 minutes/day in moderate-to-vigorous PA (MVPA) and accumulated 13334±4026 steps/day on average (Table 1). The majority (71%) meet professional guidelines of ≥60 minutes of MVPA/day for PA in children and adolescents [3].

**Table 1.** Participant characteristics, Actigraph estimates, and measures of somatic growth and maturation. All are provided in the full sample, separately by sex, and within sex by Tanner stage. Means and (standard deviations) and significant sex differences presented below.

|  | Females |  |  |  |  |  | Males |  |  |  |  |  | Full Sample |
| --- | --- | --- | --- | --- | --- | --- | --- | --- | --- | --- | --- | --- | --- |
|  | All | T 1 | T2 | T 3 | T 4 | T5 | All | T 1 | T2 | T 3 | T 4 | T5 |  |
| N | 55 | 10 | 9 | 10 | 10 | 16 | 55 | 14 | 10 | 10 | 9 | 12 | 110 |
| Age | 13.7<br>(3.1) | 10.1<br>(1.0) | 11.2<br>(1.9) | 13.1<br>(1.1) | 14.3<br>(1.1) | 17.5<br>(1.8) | 13.7<br>(3.3) | 9.9<br>(1.2) | 12.4<br>(1.0) | 13.4<br>(0.8) | 15.0<br>(1.1) | 18.3<br>(1.9) | 13.7<br>(3.2) |
| **Anthropometry** |  |  |  |  |  |  |  |  |  |  |  |  |  |
| Weight (kg) | 41.8 (10.1) | 27.7<br>(2.1) | 33.0<br>(3.4) | 44.5<br>(4.7) | 46.8<br>(4.2) | 50.7<br>(6.7) | 43.5 (15.7) | 26.7 (3.0) | 37.5<br>(8.3) | 40.1<br>(4.7) | 51.0<br>(4.9) | 65.6 (11.0) | 42.7 (13.1) |
| Height (cm) | 143.6 (9.2) | 129.2 (4.4) | 137.5 (6.3) | 147.6 (5.3) | 148.7 (3.3) | 150.3 (3.2) | 146.4<br>(15.0) | 126.6<br>(4.6) | 140.4 (6.4) | 147.5 (5.5) | 158.6 (3.2) | 164.4 (2.7) | 145.0<br>(12.4) |
| Body Fat (%) | 19.2**<br>(2.8) | 16.1<br>(1.9) | 17.8<br>(1.4) | 20.0<br>(2.9) | 21.6<br>(1.8) | 20.1<br>(2.1) | 6.6<br>(3.1) | 4.9<br>(2.2) | 6.2<br>(1.7) | 5.9<br>(1.7) | 7.3<br>(1.2) | 9.1<br>(5.0) | 13.0<br>(7.0) |
| **Actigraph** |  |  |  |  |  |  |  |  |  |  |  |  |  |
| Sedentary (min/d) | 381.6*<br>(69.1) | 294.6<br>(49.9) | 334.9<br>(35.1) | 386.7<br>(36.6) | 432.9<br>(60.0) | 430.0<br>(37.2) | 330.2<br>(83.5) | 247.1<br>(56.3) | 318.9<br>(55.7) | 331.8<br>(72.3) | 411.6<br>(77.4) | 380.4<br>(47.0) | 356.4<br>(80.4) |
| Light intensity PA (min/d) | 320.7<br>(71.7) | 395.4<br>(26.8) | 380.6<br>(36.9) | 307.7<br>(37.5) | 280.1<br>(77.9) | 268.1<br>(55.1) | 333.7<br>(79.4) | 390.4<br>(76.0) | 355.8<br>(50.6) | 363.2<br>(67.1) | 279.1<br>(58.6) | 267.1<br>(50.7) | 327.1<br>(75.5) |
| MVPA (min/d) | 63.7**<br>(32.7) | 87.2 (45.7) | 65.5 (13.4) | 67.4 (35.8) | 59.3 (30.1) | 47.0 (21.6) | 121.9<br>(45.1) | 133.6<br>(42.1) | 119.1<br>(27.3) | 104.3<br>(36.1) | 113.9<br>(73.7) | 128.7<br>(42.0) | 92.3 (48.8) |
| Steps/Day | 11464.2**<br>(3130.4) | 13371.3<br>(3091.5) | 12494.1<br>(2989.0) | 11976.6<br>(3368.2) | 10876.8<br>(2421.4) | 9585.6<br>(2652.6) | 15277.6<br>(3954.4) | 17443.8<br>(4123.0) | 15671.8<br>(3497.8) | 14692.8<br>(2881.7) | 13745.2<br>(4762.9) | 13947.9<br>(3567.2) | 13334.2<br>(4025.9) |
| **Growth and Maturation** |  |  |  |  |  |  |  |  |  |  |  |  |  |
| Height Velocity (cm/year) | 3.7<br>(2.9) | 5.5<br>(1.0) | 6.4<br>(1.0) | 6.0<br>(1.8) | 2.7<br>(1.6) | 0.2<br>(0.9) | 4.5<br>(2.2) | 4.7<br>(0.8) | 5.5<br>(1.3) | 6.4<br>(1.6) | 4.8<br>(2.7) | 1.7<br>(1.5) | 4.1<br>(2.6) |
| Grip Strength | 19.9*<br>(5.6) | 14.7<br>(2.3) | 15.8<br>(3.5) | 20.8<br>(4.8) | 21.5<br>(3.5) | 23.9<br>(5.9) | 24.9<br>(11.4) | 13.9<br>(1.8) | 18.0<br>(3.5) | 21.7<br>(4.6) | 31.6<br>(7.2) | 41.1<br>(6.4) | 22.4<br>(9.3) |
| Log DHEA (pg/ml) | 10.3<br>(1.0) | 9.4<br>(0.8) | 9.8<br>(0.9) | 10.0<br>(0.7) | 10.9<br>(0.4) | 10.8<br>(0.9) | 10.1<br>(1.0) | 9.1<br>(0.8) | 9.8<br>(0.7) | 10.1<br>(0.6) | 10.8<br>(0.9) | 10.9<br>(0.7) | 10.2<br>(1.0) |
| Log Testosterone (pg/ml) | 9.9<br>(0.8) | 9.3<br>(0.8) | 9.6<br>(0.8) | 9.8<br>(0.8) | 10.1<br>(0.3) | 10.3<br>(0.8) | 9.8<br>(0.9) | 9.0<br>(0.6) | 9.5<br>(0.7) | 10.0<br>(0.6) | 10.3<br>(0.9) | 10.6<br>(0.7) | 9.9<br>(0.9) |

Abbreviations: T=Tanner stage, MVPA: moderate to vigorous physical activity, DHEA: dehydroepiandrosterone
\*Significant sex difference, $p < 0.01$
\*\* Significant sex differences, $p < 0.001$

Self-reported activities from 24-hour recall interviews are presented in Table S2. Device-based measures of PA and sedentary were minimally impacted by attending school, as the majority of participants (60%) spent no time in school during study participation, while 24% spent one 4-hour day and 16% spent two 4-hour days in school, with no sex differences and younger children more likely to have spent time in school than older children. In the full sample, recalled wake-time was divided accordingly: sedentary leisure activities (33%); sedentary habitual/obligatory activities (22%); light intensity habitual/obligatory activities (11%); moderate to high intensity transportation (11%), habitual/obligatory activities (10%), and leisure activities (8%); followed by light intensity leisure activities (4%) and transportation (1%) with sedentary transportation (<1%) making up the smallest proportion of recalled wake-time.

### Device-measured sedentary and PA behavior by sex and age (P1 and P2)

Consistent with P1, age was positively associated with sedentary time, and negatively associated with light PA and steps/day (*p’s* < 0.001), and MVPA (*p=*0.05, Table 2a), adjusting for sex. Each year increase in age was associated with 4% more sedentary time (15 mins/day, SE=1.81), 6% less light intensity PA, 4% less MVPA, and 426 fewer steps/day (4%). Consistent with P2, males had 14% less sedentary time than females (330±84 vs. 382±69 mins/day), 91% more MVPA minutes/day(122±45 vs. 64±33 minutes/day) and accumulated 33% more steps/day (mean±SD: 15278±3954 vs. 11524±3819 steps/day), while light PA was not significantly different in males and females (Table 2). Sex differences remained significant after controlling for age (*p’s* < 0.001, Table 2a). Sex × age interactions were not significant for any PA outcome, so were not included in the reported models.

**Table 2.** Linear regression model results for sedentary time and physical activity.

| **Outcome** | **Predictor** | **Unstd.**<br>**B** | **SE** | **p-value** | **Adjusted**<br>**R<sup>2</sup>** |
| --- | --- | --- | --- | --- | --- |
| Sedentary (mins/day) | Age | 15.1 | 1.81 | <0.001 | 0.46 |
|  | Sex | -50.6 | 11.52 | <0.001 |  |
| Light intensity PA (mins/day) | Age | -15.1 | 1.78 | <0.001 | 0.41 |
|  | Sex | 12.5 | 11.36 | 0.27 |  |
| MVPA (mins/day) | Age | -2.3 | 1.18 | 0.05 | 0.37 |
|  | Sex | 58.2 | 7.6 | <0.001 |  |
| Steps/day | Age | -426 | 100 | <0.001 | 0.33 |
|  | Sex | 3780 | 646 | <0.001 |  |

### Mediation through Tanner stage (P3)

Standardized estimates from structural equation models (SEM) testing the mediating effect of Tanner stage on the associations between age and sedentary/PA outcomes are presented in Table 3 and Figure 1. The indirect effect of Tanner stage significantly mediates the associations between age and all four outcomes: sedentary mins/day (*z*=3.97, *p*<0.001), light intensity PA mins/day (*z* = -2.05, *p* = 0.04), MVPA (*z* = -2.68, *p* = 0.005) and steps/day (*z*= - 2.81, *p* = 0.005). The direct effects between age and sedentary/PA outcomes are no longer significant when accounting for the indirect effect through Tanner stage except in the MVPA model (*z* = 2.27, *p* = 0.02). The correlation matrix for SEM variables is presented in Supplementary Table S3.

**Table 3.** Mediating Effects of Tanner stage from Structural Equation Models.

| Outcome Variables | Exploratory variables | Standardized Estimate | SE | p-value |
| --- | --- | --- | --- | --- |
| Sedentary Minutes | Direct Effect - Age | -0.32 | 0.07 | 0.16 |
|  | Indirect Effect - Tanner | **0.93** | **0.23** | **<0.001** |
| Light Minutes | Direct Effect - Age | -0.08 | 0.27 | 0.78 |
|  | Indirect Effect - Tanner | **-0.57** | **0.27** | **0.04** |
| MVPA Minutes | Direct Effect - Age | **0.64** | **0.28** | **0.02** |
|  | Indirect Effect - Tanner | **-0.81** | **0.28** | **0.005** |
| Steps per day | Direct Effect - Age | 0.49 | 0.30 | 0.10 |
|  | Indirect Effect - Tanner | **-0.85** | **0.30** | **0.005** |

MVPA = Moderate to vigorous physical activity

**Figure 1a-d.**
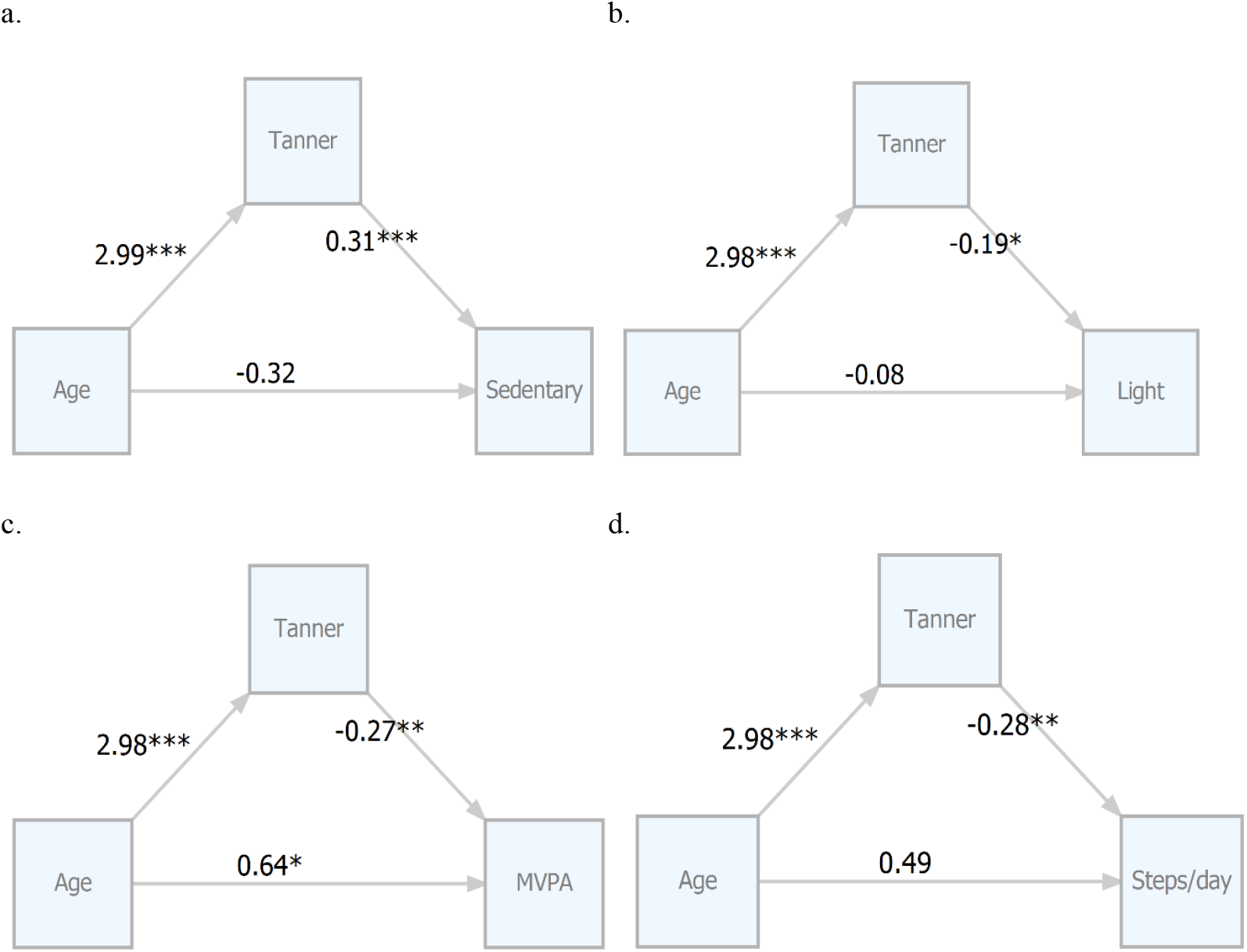
Structural equation models testing Tanner s. Standardized estimates for each model testing Tanner stage as a mediator of the relationships between age and PA outcomes (a. Sedentary time, b. Light intensity PA, c. MVPA, d. Steps/day) in the full sample. Abbreviations: MVPA moderate-to-vigorous physical activity,\**p*<0.05, \*\**p*<0.01, \*\*\**p*<0.001

The indirect effect of age on sedentary time through Tanner indicates that each year increase in age is associated with an increase of 23 sedentary minutes/day (7%). The indirect effect of age on light intensity PA through Tanner suggests that each year increase in age is associated with a decrease of 12 minutes of light intensity PA/day (4%). The direct effect of age on MVPA suggests that each year increase in age is associated with an increase of 9 minutes of MVPA/day (10%), whereas the indirect effect of age through Tanner suggests that each year increase in age is associated with a *decrease* of 11 minutes of MVPA/day (12%). The indirect effect of age on steps/day through Tanner indicated that each year increase in age is associated with a decrease of 1014 steps/day (8%). Figure 2 displays PA time and intensity within-sex by Tanner stage.

**Figure 2.**
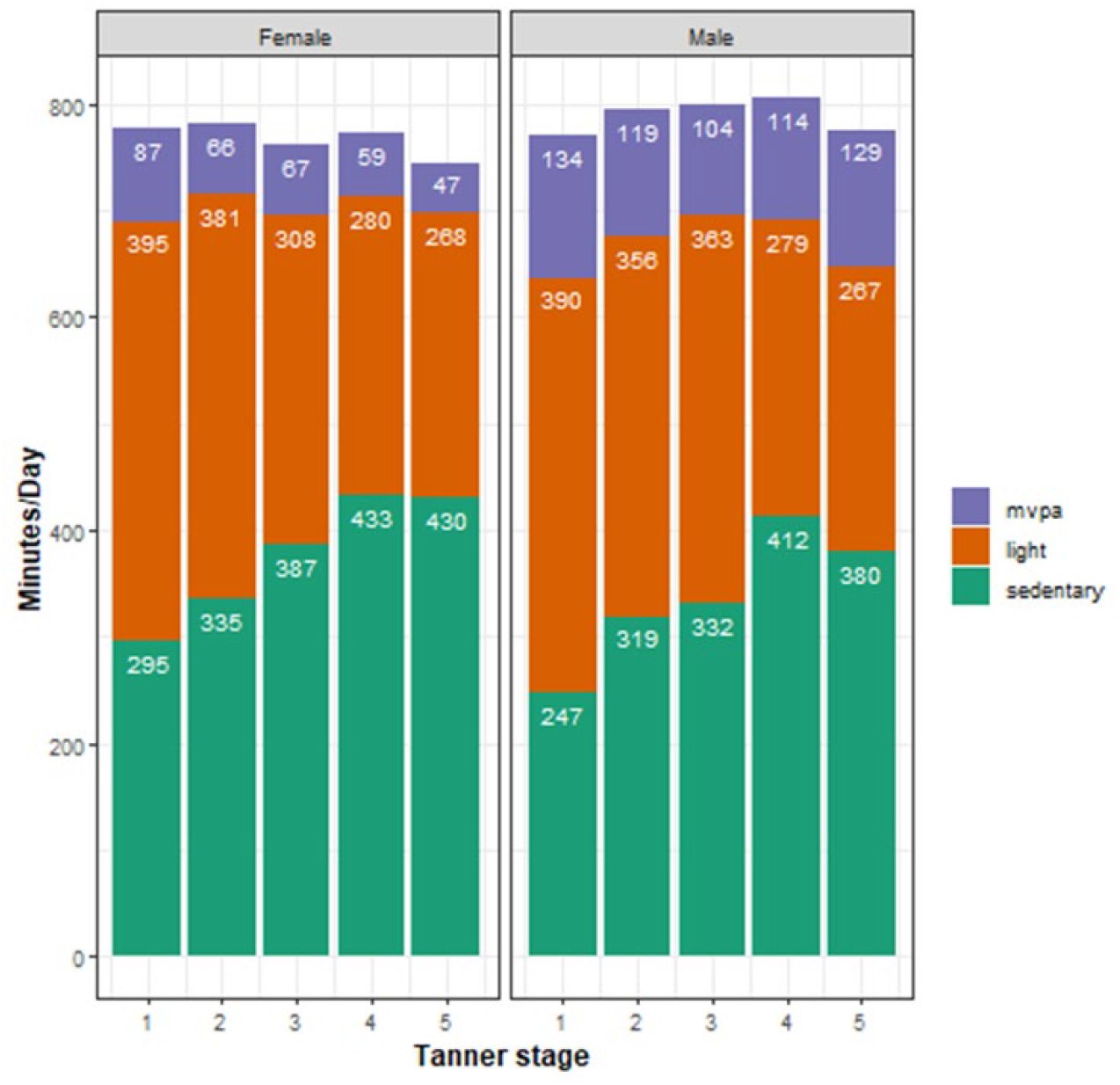
Device-measured physical activity within-sex by Tanner stage. Abbreviations: MVPA moderate-to-vigorous physical activity.

### Self-reported activity type and intensity by sex and Tanner stage

Figure 3 displays the proportions of recalled wake time spent in each activity type/intensity by Tanner stage and within-sex. Both males and females in later Tanner stages reported smaller proportions of time in leisure time MVPA (e.g., playing chase, playing with others, playing soccer) in a fairly linear pattern, which started and ended higher in males (males: 20% in Tanner 1 to 8% in Tanner 5 vs. females: 8% in Tanner 1 to <1% in Tanner 5). Similar linear patterns across Tanner stages were also observed for changes in habitual/obligatory activities (e.g., labor to produce/process food): females in later Tanner stages reported larger proportions of time in light intensity habitual/obligatory activities like food processing and washing clothes (6% in Tanner 1 to 30% in Tanner 5), whereas males in later Tanner stages reported a larger proportion of time spent in moderate-to-high intensity habitual/obligatory activities like chopping trees and accompanying others to get materials from the forest (6% in Tanner 1 to 14% in Tanner 5).

**Figure 3.**
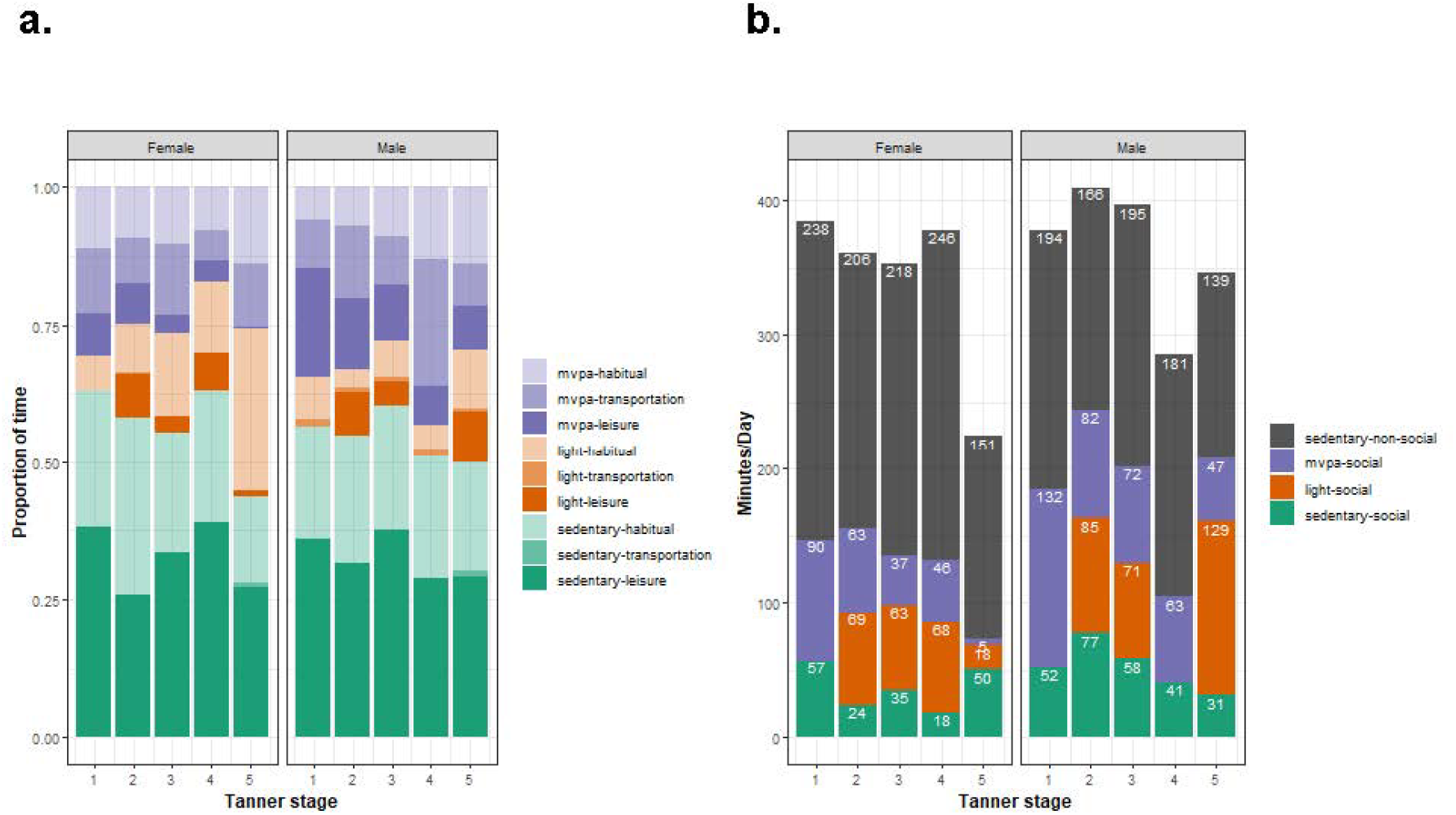
Self-reported activities from 24-hour recall interviews. Activities detailed in Table S1 are pictured within-sex and by Tanner stage. (a) includes proportions of time spent in activities classified by type and intensity, (b) illustrates time spent in leisure activities of different intensity separated by social and non-social activities. Proportion of time is included in (a) because there were individual differences in the: amount of time where activities could be reliably recalled, and time spent in sleep and activities with the medical team and anthropologists that were removed from analysis.

Females in Tanners 1-4 reported similar proportions of time in moderate-to-high habitual/obligatory activities (∼10%), and Tanner 5 females reported a similar proportion of time as males (14%). Males reported negligible light intensity habitual/obligatory activities. Three categories were fairly consistent across Tanner stages and sexes: Moderate-to-high intensity transportation (∼10%), sedentary habitual/obligatory (∼20%), and sedentary leisure (∼30%). Leisure time activities were further separated into communication-based social activities (all sedentary and light intensity), play-based social activities (all moderate to high intensity), and non-social activities (Supplemental Figure 1b). Time spent in communication-based social activities were higher among those in Tanner 2-4 than Tanner 1, while time in play-based social activities was lower in later Tanner stages and non-social sedentary activities were stable across Tanner stages.

## Discussion

Given the high prevalence of insufficient physical activity in adolescents worldwide and the increasing prevalence of childhood obesity and lifetime chronic diseases associated with inactivity, this study sought to help clarify the underlying biological and environmental factors that influence PA and sedentary time at this critical life stage. In line with our predictions, we found a significant positive association between age and sedentary time, and negative associations between age and steps/day and time spent in light intensity PA and MVPA among Tsimane children and adolescents despite their rural, non-industrialized, subsistence lifestyle. The age-activity associations did not vary by sex, and males and females had similar levels of light intensity PA. Males spent significantly less time being sedentary, almost twice as much time in MVPA, and accumulated more steps/day than females.

These findings provide an important comparison to help disentangle the effects of the environment and biology on PA during adolescence. The strikingly similar patterns observed in the Tsimane and a large body of epidemiological research conducted in post-industrialized populations, suggests that the age-activity associations and sex differences in PA/sedentary time in the transition from childhood to adulthood may be based on universal energetic tradeoffs during the pubertal transition, and hence an innate feature of the human lifespan. Tsimane children and adolescents live in an environment that is more reminiscent of those faced by our ancestors for most of human history than in post-industrialized settings. During the period of study, they did not have access to computers, video games, tablets, or smartphones; they spent very little time in formal schooling; and they are maturing into a subsistence-based economy that relies on high levels of PA to produce food through foraging and small-scale horticulture. Despite these environmental differences, older adolescents spend more time being sedentary, less time in light intensity PA and MVPA, and accumulate fewer steps/day than children.

In contrast, absolute levels of PA were much higher and the prevalence of insufficient PA is much lower in the Tsimane compared to most populations studied to date, even when comparing device-measured PA in the Tsimane to self-reported survey data that tends to *overestimate* PA. Just 29% of our sample, which had a broader age range, do not meet PA guidelines, whereas 80% of adolescents (aged 10-17 years) worldwide do not meet PA guidelines based on a recent pooled analysis of survey data [3]. Even though MVPA levels were high, Tsimane in our sample still self-reported spending most of their time in sedentary leisure and habitual activities, as well as light intensity habitual/obligatory activities, consistent with observations in Tsimane adults [38], and other indigenous populations with highly active lifestyles [42], demonstrating that sedentary behaviors are common in people of all ages, and leisure time PA is not the primary contributor to high levels of PA observed in adults in pre-Industrialized populations.

This study also helps clarify why biological maturation, rather than age alone, is a stronger predictor of PA during adolescence. We tested predictions derived from a life history framework, that associations between age and activity are explained by a wide range of anatomical and neuroendocrine changes encompassed within Tanner stages by testing mediational path models in SEM with indirect paths between age and activity through Tanner stage. Our mediational results demonstrate that Tanner stage explains the relationships between age and sedentary time, MVPA mins/day, steps/day, and to a lesser degree, light intensity PA. In all models but MVPA, the inclusion of a mediational path through Tanner eliminated the significant relationship between age and PA.

The current findings do not support the hypothesis that adolescents become more sedentary as a form of increasing health-risk behaviors in response to the psychosocial stress of puberty because physical inactivity is not a health-risk behavior for the Tsimane. Instead, our findings support our primary hypothesis that the energetic needs for growth, maturation, and reproduction are heightened during adolescence, and so trade-off against the energy available for PA, regardless of the environmental context. Another study of device-measured PA in adolescent Yucatec Maya agriculturalists also supports the hypothesis that energy for growth competes with energy available for PA [43]. In that study, observational physical activity measures and anthropometry were measured over a 20-year span during which a school and mechanized technologies were introduced to a rural community, but diet (energy availability) was minimally changed. PA level declined substantially, while height, weight, and body fat increased. It could also be that the importance of building embodied social capital through communication-based social activities becomes increasingly important during reproductive maturation [44]. The self-report data revealed that lower intensity communication-based leisure activities were more prevalent in Tanner stages 2-5 than Tanner 1, while play-based social leisure-time activities. Time spent in non-social leisure activities were similar across individuals in all Tanner stages. Obligatory and work-related activities were also reported more frequently in more mature Tanner stages.

Of note, older Tsimane males in our sample, classified as Tanner 4 and 5, had higher mean MVPA mins/day than those in Tanner stages 2 and 3, but lower than Tanner 1 (Table 1), which coincides with the stages where mean height velocity was the highest. This contrasts with long-term longitudinal studies in the UK using device-measured PA and Canada using self-reported PA which found that PA in adulthood remained lower after the adolescent decrease in activity [13,14]. Although the direction of this effect is opposite in terms of the association between age/Tanner and PA in older adolescents in different contexts, it also highlights a similarity in the importance of economic productivity for influencing PA in older adolescents. In non-industrialized contexts, food and economic production require high levels of PA; whereas in post-industrialized contexts, food production requires minimal physical effort and future economic productivity can benefit from more sedentary behaviors (i.e., more schooling). Indeed, Tsimane males in later Tanner stages reported doing more moderate to high intensity activities like chopping trees, clearing foliage, getting materials from the forest, than those in earlier Tanner stages, and less higher intensity leisure activities, like playing soccer or playing chase. Similarly, Tanner 5 females reported spending <1% of time in higher intensity leisure activities, but a larger proportion of time in light *and* moderate-to-high intensity habitual/obligatory activities, like tending children, washing clothes, removing rice from the hull, and retrieving firewood. In line with a study of Hadza children and juveniles [45], sexual division of labor appears in middle-childhood and can be seen even in those classified as Tanner 1, with habitual/obligatory activities classified as being lighter intensity reported more among females, whereas moderate-to-high intensity obligatory activities were reported more frequently among males.

In the current study, Tanner stage serves as a proxy for a variety of endocrine and somatic changes that characterize the transition from childhood to adulthood. Tanner stage was significantly associated with DHEA, testosterone, height velocity, strength, and body fat in males and females in ways that are consistent with the existing literature conducted in post-industrialized populations (Table S2) [46,47]. Notably, sex differences in body fat are evident in Tanner 1, and body fat was still more strongly associated with Tanner and all hormonal and somatic measures of growth and maturation in females. In contrast, grip strength was more strongly associated with Tanner in males compared to females. The association between Tanner and height velocity in males was relatively weak. This is likely driven by correlations only testing the linear relationship between variables, whereas the means of height velocity by Tanner stage suggest that there is a quadratic relationship between height velocity and Tanner stage that is particularly evident in males because of the later timing of maximum height velocity in Tanner 3 rather than Tanner 2 in females (Table 1).

Given the Tsimane are a food-limited population with high pathogen burden, natural fertility, and earlier age of first birth [41,48,49], the energetic trade-offs between growth/maturation/reproduction and energy available for PA may be more intense than for those in energy-rich, post-industrialized populations where food availability is much higher, and the immune system burden are much lower. However, this straightforward prediction is complicated by the fact that subsistence lifestyles require high levels of PA. Direct comparisons between our data and the existing literature are difficult, as most studies use self-report PA [9], or a different measure of biological age or metric for device-measured PA by Tanner [14,21]. A larger, longitudinal study among Tsimane with a broader range of energetic condition and access to food can help clarify if this energetic condition moderates the effects of Tanner stage on physical activity.

There is some evidence that the Tsimane exhibit steeper trade-offs between growth/maturation/reproduction and activity. The highest mean height velocity in our sample was the same for males and females (6.4 cm/year), which is lower than values observed in the US which also vary by sex (females: 9cm/year, males: 10 cm/year) [37]. In addition, trade-offs between reproductive efforts and energy for PA may be more severe for Tsimane females compared to Industrialized populations. Tsimane males in our sample accumulated 33% more steps/day than females; whereas a study in Australian males reported only 19% more steps than females measured by pedometer, although their age range was smaller, 8-12 [50]. Tsimane males in the current study also have 91% more MVPA minutes/day than females, whereas males in the US between the ages of 6-19 had 72% more MVPA minutes/day, though it should be noted that in ages 6-11 boys had 162% more MVPA minutes/day than girls [5]. Portuguese males between 13-14 have 39% more MVPA minutes/day than females, 13 year old Canadian males have 31% more MVPA than females [51], and ∼12 year old males in the UK have 56% higher MVPA minutes/day than females [21]. Notably, the global study of self-report PA [3], the prevalence of insufficient PA was higher among adolescents in low-income countries than those in middle to high income countries, while this pattern is reversed in adults [2]. Overall, adolescents in post-industrialized countries exhibit low levels of PA levels, despite more access to food and lower immune system burden.

In terms of the underlying biology, this pattern suggests that additional energy availability in Industrialized environments is prioritized for skeletal growth, energy storage, and higher levels of reproductive hormone over increased motivation to expend energy in PA.

The primary limitation of this study is that it was cross-sectional. A longitudinal design would provide a clearer picture of the associations between growth and maturation on PA/inactivity. In addition, our sample size within each Tanner stage was small, and all data were collected in one season when school was not in session, so we cannot generalize the PA levels we found to those experienced year-round, or while school is in session. However, an advantage of this design is that our PA estimates were not affected by seasonality, nor by school-age children attending school and older adolescents not attending school. In addition, Tanner stage captures a constellation of changes that occur during the transition from childhood to adulthood. Several changes are co-occurring at this life stage, and likely affect each other, as well as behavior. Future work aiming to tease apart the differential effects would benefit from including additional measures, particularly hormones related to growth, (e.g., IGF-1) and energetic condition (ghrelin, leptin, C-peptide). A complete life history model would also include measures of diet, energetic condition, as well as immune markers, to capture the roles that these factors play in shaping the energy available for PA in children and adolescents. However, this analysis is meant to represent an important first step toward clarifying the relationships between age, maturation, and PA in adolescence, and these limitations are balanced with several other strengths. This study was conducted in a unique population living a vastly different lifestyle in a vastly different environment than populations typically studied in the epidemiology or kinesiology literature. In addition, we used device-measured estimates of PA time and intensity, as well as self-reports to capture the types of activities Tsimane children and adolescents engage in. We also included Tanner assessments and validated them with hormone levels and markers of somatic growth and maturation and included a wide age-range of participants to provide a better picture of PA across the transition from childhood to adulthood.

Taken together with the existing literature, our findings raise important considerations for improving public health in energy-rich, industrialized environments that are mismatched to environments faced by humans for the vast majority of our evolutionary past. Adolescence has emerged as a critical window to intervene on exercise and PA engagement, but our findings, and those of several others in post-industrialized settings, suggest that the limited time and resources for public health interventions may be more effective if targeting increasing PA in early to mid-childhood. Interventions in adolescents may be more effective if they focus on breaking up sedentary time, and limiting reductions in PA, rather than increasing PA participation. Adolescents may also be more inherently drawn to lower intensity activities that provide opportunities for social engagement. Sex-specific interventions may also be needed once adrenarche and puberty lead to sexual dimorphism in the absolute and relative costs and benefits of PA. The most effective intervention may require modifying our environments to incorporate more PA in our habitual and obligatory activities, providing safe and enjoyable opportunities for active transportation, rather than relying exclusively on exercise and sports participation to reach PA levels consistent with guidelines. Negative associations between reproductive maturation and PA also introduces additional consequences for energy-rich populations where maturation is occurring at earlier ages. Shorter childhood means fewer years are spent where the costs of PA participation are low. Additionally, earlier maturation may lead to a disconnect between physical, brain, and social development that make it more difficult to navigate and can lead to less health promoting behavior.

Life history theory provides a useful framework for understanding the complex set of factors that influence PA in adolescence and across the lifespan. Absolute levels of PA are strongly influenced by the amount of obligatory PA in a given environment. Cross-cultural consistencies in PA patterns observed in Tsimane adolescents and those from post-industrialized countries suggests that a coordinated system has evolved that responds to intensifying energetic needs for growth and maturation during adolescence by prioritizing energetic effort toward growth and maturation over leisure-time PA, irrespective of energy stores, or cues of energy abundance in the environment. Maturation and cognitive development may also include an increased interest in building social relationships through communication-based social activities that are lower intensity, and a decreased interest in higher intensity, play-based social activities. The constellation of life-history shifts that are simultaneously occurring: increased stature, muscle and fat deposition, and social skill development each impact PA. Growth, reproductive maturation, and reproduction place fundamental constraints on the energy, time, and motivation for PA that warrant consideration in future PA research and intervention development.

## Materials and Methods

### Experimental design

Data for this cross-sectional study were collected among Tsimane children and adolescents from four adjacent villages located along the Maniqui River in the summer of 2014 in collaboration with the Tsimane Health and Life History Project [48]. To encompass the transition from childhood to adulthood, a wide age range of participants (ages 7-22) was recruited, in line with a broader conceptualization of adolescence that extends the upper limit to align more closely with brain development [52]. Tsimane children and younger adolescents play with others in neighboring families, assist in caring for younger siblings and food production, and spend limited time in formal schooling compared to children in post-industrialized populations. Older Tsimane adolescents in this age range are typically married with children, and produce food, but typically reside in family clusters and are still reliant on their parents and grandparents for additional food production and allocare [53].

Participants were recruited during their annual medical exam with the THLHP medical team by AC and PH. All individuals between the ages of 7-22 who attended their medical exam were invited to participate (n=126, 37% of individuals in this age range resident in these communities in 2014).

Participation lasted 3-4 days allowing inclusion of all eligible participants from each community during the medical team visit (1-3 weeks). On Day 1 participants and parents provided informed consent/assent, completed a structured interview and had anthropometric and strength measurements taken (detailed below). Participants wore ActiGraph wGT3X+ accelerometers on their hip for the duration of study participation. On days 2 and 3, participants provided first void urine samples to measure urinary hormones (detailed below). On days 2, 3, and 4 (where possible) participants completed 24-hour physical activity recall interviews while concurrently examining Actigraph data on a computer screen to validate wear-time and aid in recall. Small gifts were provided to remunerate participants for each day of study participation including pens and notebooks, backpacks, soccer jerseys, sardines, and oil.

All study methods were approved by the Institutional Review Board (IRB) at the University of New Mexico (HRRC # 07–157). Informed consent was established at three levels: (1) the Tsimane governing council; (2) village leadership; and (3) study participants. Informed assent was established with parents/guardians of minors.

### Participant Characteristics

A total of 110 Tsimane children and adolescents (mean±SD age = 13.7±3.2 y, 50% female) were sampled from four villages (Figure 4). Table 1 summarizes sample characteristics for all variables of interest in the full sample, separately by sex, and within-sex by Tanner stage. Males and females were similar with respect to mean age, weight, height, DHEA, testosterone, and distribution across Tanner stages, *Χ*^*2*^(4, n=110) = 1.34, *p*=0.85. Males have significantly higher grip strength, non-significantly (*ns)* higher mean height velocity (4.5 ± 2.2 vs. 3.7 ± 2.9 cm/year, *p*=0.09); while females had significantly higher mean body fat. The highest mean height velocity across Tanner stages was the same in males and females (6.4 cm/year) and was observed in Tanner 2 females and Tanner 3 males.

**Figure 4.**
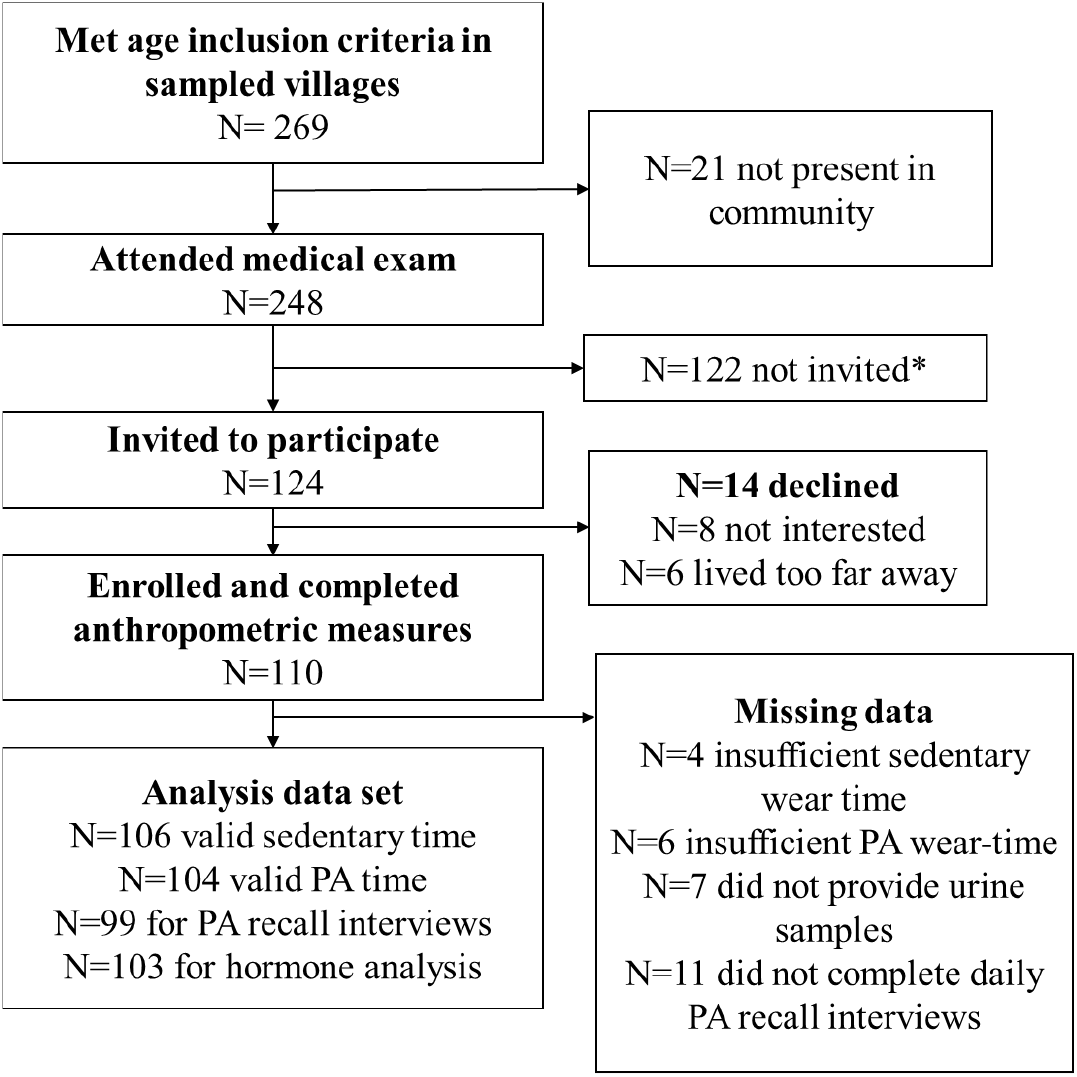
STROBE diagram detailing sample selection and data collection. Abbreviations: PA – physical activity. *Individuals who met the inclusion criteria and attended a medical exam were invited depending on the availability of accelerometers. When accelerometers were available, invitations were prioritized for individuals who were co-resident in the same household (to reduce logistical costs), and when inclusion improved sample coverage across age-sex categories. This resulted in 124 individuals being invited, and 122 not being invited.

### Measures

#### Age and Anthropometry

Age was obtained from census data collected by the Tsimane Health and Life History Project (THLHP) over the past 20 years. Given the young age range of the current sample, dates of birth are very accurate. Height was measured to the nearest cm with a portable stadiometer (Seca 213) and weight with a digital scale (Tanita BC-1500). Body fat was estimated using the Jackson & Pollack 3-measurement skinfolds taken in triplicate using FatTrack®II Digital Body Fat Calipers (AccuFitness). Grip strength was measured taking the average of three measurements [54] using a hydraulic dynamometer (Baseline, TN).

#### Free-living physical activity and sedentary time

ActiGraph wGT3X+ accelerometers were used to estimate PA/sedentary time and intensity. Actigraphs were initialized with a 30hz sampling rate. Participants were fitted with the device on the hip, instructed on the proper positioning and to only remove the device when submerging themselves in water. Daily 24-hour activity recall interviews were completed to assess activity types among participants who lived within a reasonable distance from the mobile medical camp and could return daily. To aid participants’ recall of the previous day and validate Actigraph wear-time in real-time, investigators and participants viewed Actigraph data together on a laptop screen while completing the interview. Completed files were downloaded using Actilife software with the normal filter into 60 second epochs and converted to csv tables. Data were processed in R (v4.1). Wear-time was additionally assessed using a criterion of 60 consecutive minutes of zero-counts with an allowance of 1-2 minutes of counts to 100 [55]. Epochs were delineated into periods of sedentary, light and MVPA activity-levels with an algorithm developed in Brazilian adolescents using vector magnitude counts [56]. Sixty-second activity data were summed into person-day summaries including separating the 12-hour periods from 7am to 7pm and 7pm to 7am. Person-day summaries were averaged into individual daily estimates of activity. Participants were included in analysis if there was at least 10 hours of valid wear time between 7am and 7pm. Reported daily estimates for light intensity, MVPA, and steps/day were based on a 24-hour day from 7am until the following morning at 7am; while sedentary activity was based on a 12-hour period from 7am-7pm to separate daytime sedentary behavior from sleep. For daytime sedentary behavior, person-day level data were included if there were at least 9 hours of valid wear time between 7am and 7pm. Of the initial 110 enrolled in the accelerometry protocol, four did not meet the criteria of at least 1 valid day daytime sedentary behavior and an additional two participants did not meet the valid wear-time criteria for PA (Figure 4). The remaining participants provided 1.9 days (SD=0.4) of data.

Daily 24-hour activity recall interviews were completed among n=99 participants, covering an average of 3.8 days (SD=0.06), 15.6 hours per day (SD=3.0), and 3601 minutes per participant (SD=962). An average of 37 activities (SD= 9.1) were recorded per participant, and a total of 62 activities were recorded overall. These 62 activities were coded using time allocation codes for the Tsimane project used previously [57]. Activities were grouped into 9 bins along two dimensions: intensity (sedentary, light, moderate to high) and type (transportation, habitual/obligatory, and leisure) by AC and HD and modified until consensus was reached among all authors who have completed fieldwork with the Tsimane (Table S2). Sleep was not included in results and accounted for 43.3% of total recall time. Time-periods when participants couldn’t recall what they were doing for a given time-period, and activities involving medical exams or interaction with anthropologists were also excluded, accounting for 8.9% of time captured by the interviews.

#### Tanner Stage

Tanner staging is considered the gold standard for tracking the development and sequence of secondary sex characteristics throughout puberty. It is typically evaluated on a scale from 1-5 by a physician through clinical examination of secondary sex characteristics such as pubic hair growth and breast/genital development [58]. However, it is challenging to perform in some settings because of the sensitivity of the assessment and the need for privacy – particularly in field settings where direct examinations, interviews, or pictorial representations of breasts and genitals are not culturally appropriate, as in the Tsimane field setting. Thus, Tanner stage 1-5 was assessed by two authors (AC and PH) independently based on secondary sexual characteristics visible through clothing, including breast development and underarm hair, as well as menarche, lactation status and reproductive history obtained from THLHP medical records. Facial/underarm hair, muscular development, jaw shape, and voice change served as additional maturation cues in males. Inter-rater reliability was high, with discrepancies reflecting only ±1 stage difference that were resolved through discussion between researchers. Tanner stage assessments were then validated with measures of growth and reproductive maturation.

#### Urinary Hormone Assays

First morning void urine specimens were collected by participants on two consecutive days, and frozen in liquid nitrogen within several hours of specimen collection. They were stored for up to one month in liquid nitrogen, before being transferred on dry ice to the US where they were stored at -80C for two years before analyses. After arrival, specimens were thawed, specific gravity was measured with a refractometer (Atago, Inc), and urine specimens were analyzed in duplicate via enzyme immunoassay for DHEA (Enzo Life Sciences, ADI-901-093) and testosterone (R156/7[59]). All specimens were run on the first freeze thaw, and results were specific gravity corrected [60]. See Supplemental Information for a detailed description DHEA and testosterone during maturation.

#### Height Velocity

To compute height velocity in 2014, height measurements collected on a previous round by the THLHP were used to estimate height velocity. Prior measurements were taken between 1-4 years prior to 2014, with the majority (70%) taken 2 years prior to the study. Height velocity was calculated as:

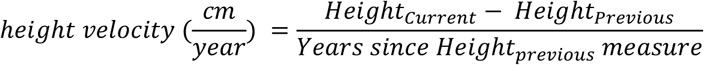

### Statistical analysis

Analyses were performed in R version 4.1. Sex differences in participant characteristics were assessed using independent sample t-tests for continuous variables and Chi-square tests for distribution across Tanner stages. Spearman correlations were tested to validate Tanner stage with log transformed hormone levels (DHEA and testosterone which were both skewed) and measures of somatic growth and maturation (height velocity, grip strength, and body fat). Non-parametric tests were used because Tanner is ordinal, and body fat in males violated normality assumptions [61]. To test P1 and P2, separate linear regression models were run for each activity outcome (sedentary time, light intensity PA, MVPA, steps/day) with age, sex, and the age x sex interaction included as independent variables. To test the mediating role of Tanner on age-sedentary/PA associations (P3), structural equation models (SEM) were fit in the lavaan package version 0.6-11 using listwise deletion for participants missing sedentary (n=4) or PA outcome data (n=6). Diagonally weighted least squares estimation was used to account for Tanner being ordinal as opposed to continuous. Age and outcome variables were standardized to facilitate model convergence given the vastly different scales of the variables. Categorization and visual representation of self-reported PA was performed in R 2022.02.0 using the ggplot2 and lmer packages. All datasets generated during and/or analyzed during the current study are available from the corresponding author on reasonable request.

### Validity of Tanner Stage with Hormone Levels and Somatic Maturation Markers

Estimates of the within-sex Spearman correlations between Tanner stage assessment and DHEA, testosterone, height velocity, grip strength and body fat are presented in Supplemental Table 1. Tanner stage assessments were significantly correlated with all these variables in both sexes (*p*’s < 0.01), with coefficients in females ranging from *r* = 0.46 for testosterone to *r* = -0.79 for height velocity and in males from *r* = -0.38 for height velocity to *r* = 0.92 for grip strength.

## Data Availability

All relevant computer code for variable definitions and statistical analysis will be downloadable from the following GitHub repository: https://github.com/anncwell/tsimane-youth-activity that will be made public upon approval of the manuscript for publication. Individual-level data are stored in the Tsimane Health and Life History Project (THLHP) Data Repository, and are available through restricted access for ethical reasons. THLHPs highest priority is the safeguarding of human subjects and minimization of risk to study participants. The THLHP adheres to the CARE Principles for Indigenous Data Governance, which assure that the Tsimane: 1) have sovereignty over how data are shared; 2) are the primary gatekeepers determining ethical use; 3) are actively engaged in the data generation; and 4) derive benefit from data generated and shared use whenever possible. The THLHP is also committed to the FAIR Principles to facilitate data use. Requests for individual-level data should take the form of an application that minimally details the exact uses of the data and the research questions to be addressed, procedures that will be employed for data security and individual privacy, potential benefits to the study communities, and procedures for assessing and minimizing stigmatizing interpretations of the research results (see the following webpage for links to the data sharing policy and data request forms: https://tsimane.anth.ucsb.edu/data.html). Requests for individual-level data will require institutional IRB approval (even if exempt) and will be reviewed by an Advisory Council composed of tribal leaders, tribal community members, Bolivian scientists, and the THLHP leadership. The study authors and the Tsimane leadership are committed to open science and are available to assist interested investigators in preparing data access requests.

## Acknowledgments

We are extraordinarily grateful to our Tsimane participants and anthropologists who work with the THLHP, particularly Chichi, Agustina Bani Cuata, Emiliana Cayuba Claros, and Neisa Durbano Hista, as well as the THLHP medical team, Matt Schwartz, and Adrian Jäggi, without whom this study would not have been possible. We thank Angela Bryan, who provided guidance on the SEM analyses. AC and PH also thank their daughter, Josephine, who braved the Bolivian Amazon and celebrated her 3^rd^ birthday in the field while her parents conducted this study.

## Funding

The Tsimane Health and Life History Project (THLHP) was supported by: NIH/NIA grants R01AG024119, R56AG024119, and P01AG022500.

AC acknowledges funding from the NIH K01 HL143039 that provided funding while she wrote this manuscript.

PLH was supported by an Omidyar Fellowship from the Santa Fe Institute during the data collection for this study.

JS acknowledges IAST funding from the French National Research Agency (ANR) under the Investments for the Future (Investissements d’Avenir) program, grant ANR-17-EURE-0010.

## Author contributions

Conceptualization: AC, PH

Methodology: AC, PH, MG, JS, HK

Investigation: AC, PH

Lab assays: BT

Data processing/analysis/visualization: DC, AC, HD

Supervision: HK, MG

Writing—original draft: AC

Writing—review & editing: All authors

## Competing interests

We have no competing interests to disclose.

## Data and materials availability

All relevant computer code for variable definitions and statistical analysis will be downloadable from the following GitHub repository: https://github.com/anncwell/tsimane-youth-activity that will be made public upon approval of the manuscript for publication. Individual-level data are stored in the Tsimane Health and Life History Project (THLHP) Data Repository, and are available through restricted access for ethical reasons. THLHP™’s highest priority is the safeguarding of human subjects and minimization of risk to study participants. The THLHP adheres to the CARE Principles for Indigenous Data Governance, which assure that the Tsimane: 1) have sovereignty over how data are shared; 2) are the primary gatekeepers determining ethical use; 3) are actively engaged in the data generation; and 4) derive benefit from data generated and shared use whenever possible. The THLHP is also committed to the FAIR Principles to facilitate data use. Requests for individual-level data should take the form of an application that minimally details the exact uses of the data and the research questions to be addressed, procedures that will be employed for data security and individual privacy, potential benefits to the study communities, and procedures for assessing and minimizing stigmatizing interpretations of the research results (see the following webpage for links to the data sharing policy and data request forms: https://tsimane.anth.ucsb.edu/data.html). Requests for individual-level data will require institutional IRB approval (even if exempt) and will be reviewed by an Advisory Council composed of tribal leaders, tribal community members, Bolivian scientists, and the THLHP leadership. The study authors and the Tsimane leadership are committed to open science and are available to assist interested investigators in preparing data access requests.

## Supplementary Materials for

### Supplementary Text

#### Methods

Levels of dehydroepiandrosterone (DHEA) and testosterone were used to help validate Tanner stage assessments. Pubertal maturation consists of two distinct and overlapping components, adrenarche and gonadarche. Adrenarche involves the reactivation of the adrenal glands, dormant since infancy, and is marked by adrenal production of DHEA (*1*). In existing research in post-industrialized populations, adrenarche typically occurs around ages 6-8 in girls and 7-9 in boys, and DHEA levels continue to rise during gonadarche and into the 3rd decade of life (*1*). DHEA is one of the hormones responsible for pubic hair development (once concentrations reach levels high enough to meet the sensitivity of the target tissue at the hair follicles) as well as a neurosteroid that may contribute to behavioral and psychological changes during pubertal maturation. Testosterone increases during puberty across Tanner stages, as plays important roles in bone mass density (*2*), muscle mass (*3*), voice pitch (*4*), and brain morphology (*5*).

**Table S1.** Activity categories. All activities self-reported from 24-hour recall interviews were categorization with intensity and type as agreed upon by all authors through group consensus.

|  | Sedentary | Light Intensity | Moderate to High Intensity |
| --- | --- | --- | --- |
| **Transportation** | Travel by motorized canoe<br>Travel by vehicle | Go out looking | Walking<br>Running for transportation<br>Travel by unmotorized canoe |
| **Habitual/Obligatory** | Eat<br>School | Fishing (hook and line/barbasco)<br>Garden labor (burn, plant)<br>Harvest rice<br>Food processing/cooking<br>Domestic animal care<br>Tool manufacture/maintenance<br>Weaving basket fan, mat, sari<br>Cleaning/washing clothes<br>Bathe<br>Tending children (not holding)<br>Visit merchant | Accompanying, hunting<br>Accompanying to get materials from the forest<br>Hunt<br>Fishing with net/bow and arrow<br>Garden labor (chop trees, clear foliage with machete)<br>Remove rice from hull<br>Foraging<br>Get water<br>Get firewood<br>Clear yard, soccer field, trail, etc. with machete<br>Carrying objects (aside from water, firewood)<br>Getting materials from the forest to manufacture something (e.g., house) |
| **Leisure** | At home<br>Listen to music/radio<br>Read/write, homework<br>Lying down<br>Sit<br>Nap<br>Go out visitng/talk social<br>Watch tv* | Party (general) | Dance (social)<br>Play (general, chase, with others)<br>Play sports (soccer, volleyball) |

\*\*Light intensity was cautiously assigned to non-sedentary behaviors where specific details and techniques were not provided by interviewee
\* There was one household where tv was watched by several participants, and one instance where the mobile medial team and anthropologists showed a movie on a computer screen during participation.

**Table S2.** Correlation matrix of Tanner stage assessments and somatic growth and maturation variables.

|  | 1 | 2 | 3 | 4 | 5 | 6 |
| --- | --- | --- | --- | --- | --- | --- |
| 1. Tanner | -- | **0.697\*\*** | **0.667\*\*** | **-0.376\*** | **0.921\*\*** | **0.484\*\*** |
| 2. DHEA-S | **0.581\*\*** | -- | **0.792\*\*** | -0.222 | **0.701\*\*** | 0.279 |
| 3. Testosterone | **0.459\*\*** | **0.711\*\*** | -- | -0.22 | **0.722\*\*** | 0.247 |
| 4. Height velocity | **-0.786\*\*** | **-0.525\*\*** | **-0.384\*** | -- | **-0.29** | -0.144 |
| 5. Grip strength | **0.627\*\*** | **0.493\*\*** | **0.38\*** | **-0.451\*\*** | -- | **0.462\*\*** |
| 6. Body fat (%) | **0.617\*\*** | **0.588\*\*** | **0.323** | **-0.514\*\*** | **0.389\*** | -- |

**Table S3.** Spearman correlation matrix of variables in mediation models.

|  | 1 | 2 | 3 | 4 | 5 | 6 |
| --- | --- | --- | --- | --- | --- | --- |
| 1. Age | -- | -- | -- | -- | -- | -- |
| 2. Tanner | **0.922\*\*** | -- | -- | -- | -- | -- |
| 3. Sedentary (mins/day) | **0.648\*\*** | **0.698\*\*** | -- | -- | -- | -- |
| 4. Light intensity PA (mins/day) | **-0.662\*\*** | **-0.675\*\*** | **-0.815\*\*** | -- | -- | -- |
| 5. MVPA (mins/day) | -0.189 | **-0.251\*** | **-0.523\*\*** | 0.161 | -- | -- |
| 6. Steps/day | **-0.346\*\*** | **-0.391\*\*** | **-0.562\*\*** | **0.411\*\*** | **0.767\*\*** | -- |

## Notes

### Competing Interest Statement

The authors have declared no competing interest.

### Author Declarations

Ethics committee/IRB of University of New Mexico gave ethical approval for this work

